# SARS-CoV-2 Antibody Lateral Flow Assay for antibody prevalence studies following vaccine roll out: a Diagnostic Accuracy Study

**DOI:** 10.1101/2021.07.14.21260488

**Authors:** Alexandra Cann, Candice Clarke, Jonathan Brown, Tina Thomson, Maria Prendecki, Maya Moshe, Anjna Badhan, Paul Elliott, Ara Darzi, Steven Riley, Deborah Ashby, Michelle Willicombe, Peter Kelleher, Paul Randell, Helen Ward, Wendy S Barclay, Graham Cooke

**Author notes:** contributed equally to this work (Alexandra Cann, Candice Clarke, Jonathan Brown).

## Abstract

**Background:** Lateral flow immunoassays (LFIAs) have the potential to deliver affordable, large scale antibody testing and provide rapid results without the support of central laboratories. As part of the development of the REACT programme extensive evaluation of LFIA performance was undertaken with individuals following natural infection. Here we assess the performance of the selected LFIA to detect antibody responses in individuals who have received at least one dose of SARS-CoV-2 vaccine.

**Methods:** This is a prospective diagnostic accuracy study.

*Setting:* Sampling was carried out at renal outpatient clinic and healthcare worker testing sites at Imperial College London NHS Trust. Laboratory analyses were performed across Imperial College London sites and university facilities.

*Participants:* Two cohorts of patients were recruited; the first was a cohort of 108 renal transplant patients attending clinic following SARS-CoV-2 vaccine booster, the second cohort comprised 40 healthcare workers attending for first SARS-CoV-2 vaccination, and 21 day follow up. A total of 186 paired samples were collected.

*Interventions:* During the participants visit, capillary blood samples were analysed on LFIA device, while paired venous sampling was sent for serological assessment of antibodies to the spike protein (anti-S) antibodies. Anti-S IgG were detected using the Abbott Architect SARS-CoV-2 IgG Quant II CMIA.

*Main outcome measures:* The accuracy of Fortress LFIA in detecting IgG antibodies to SARS-CoV-2 compared to anti-spike protein detection on Abbott Assay.

**Results:** Using the threshold value for positivity on serological testing of ≥7.10 BAU/ml, the overall performance of the test produces an estimate of sensitivity of 91.94% (95% CI 85.67% to 96.06%) and specificity of 93.55% (95% CI 84.30% to 98.21%) using the Abbott assay as reference standard.

**Conclusions:** Fortress LFIA performs well in the detection of antibody responses for intended purpose of population level surveys, but does not meet criteria for individual testing.

## Introduction

As vaccination programmes for Covid-19 are rolled out worldwide, population antibody testing is useful in monitoring immune responses to vaccinations, informing discussion and decisions about booster doses, and assessing levels of potential protective immunity in the population (1).

Lateral flow immunoassays (LFIAs) have the potential to deliver affordable, large scale testing of individuals and provide rapid results without the support of central laboratories. This approach has been used across England in the REACT2 (REal time Assessment of Community Transmission 2) to estimate the number of infections during the first wave (2), monitor the decline in antibody positivity over time (3) and assess population antibody prevalence following vaccine roll-out.

Prior to the scale up of antibody testing for surveillance, extensive clinical and laboratory evaluation of diagnostic accuracy following natural infection was performed on a range of LFIA antibody tests (4,5). The test selected (Fortress, Northern Ireland) detects antibody against the spike protein of the virus (contained in all licensed vaccines) and would therefore be expected to detect vaccine induced antibody responses. This study examined the accuracy of the Fortress LFIA device in detecting antibodies in two cohorts of vaccinated individuals and explored the relationship between LFIA results and viral neutralisation.

## Methods

This was a prospective diagnostic accuracy study conducted between 23^rd^ December 2020 and 26^th^ May 2021.

### Cohort 1: Renal Transplant Cohort

#### Participants

Participants were recruited between 9^th^ June 2021 and 26^th^ May 2021. 108 renal transplant recipients who were attending clinic 28 days (range -7 to +14) following a second dose of a SARS-CoV-2 vaccine, (either BNT162b2 Pfizer-BioNTech or ChAdOx1 Oxford-AstraZeneca). Participants underwent a capillary blood draw for immediate testing on the LFIA device and, at the same time, gave a venous blood samples for later serology testing. Clinical characteristics were obtained from electronic medical records. All patients provided written informed consent.

#### Lateral flow immunoassay testing

Participants were supplied with an LFIA testing kit as used in the REACT home testing programme (6). The LFIA (Fortress, NI) detects IgG and IgM to the S1 protein. Participants were also provided with verbal instructions on how to use the test by a member of the research team, prior to performing self-testing, with support provided where necessary. The LFIA result was assessed independently by two observers. The results were reported by the colour intensity of the IgG band, and documented as either a positive or negative result.

#### Serological testing

Serological assessment was performed on the Abbott Architect SARS-CoV-2 IgG Quant II CMIA which reports a quantitative anti-Spike antibody titre. The threshold value for positivity of 7.10 binding antibody units (BAU)/ml.

### Cohort 2: Healthcare Worker Cohort

#### Participants

Participants were recruited between 20^th^ December 2020 and 31^st^ January 2021. 39 healthcare workers were consented when attending for first vaccination with BNT162b2 Pfizer-BioNTech. 38 of these participants, had repeat samples taken at 21 days post vaccination and 1 further participant had samples taken at 21 days who had not had day 0 samples. In total there were 40 participants. Data was collected on age and gender. Medical records of participants were not accessed for this cohort.

#### Lateral Flow Immunoassay Testing

Participants were supported in capillary blood draw for use with the Fortress LFIA devices as described above. Results were reviewed and recorded by the study team.

#### Serological testing

Spike protein antibodies (anti-S) were detected using the Abbott Architect SARS-CoV-2 IgG Quant II CMIA. At the time of the study, quantitative antibody titres were reported in AU/ml. To allow combination with cohort 1 data, these were converted to BAU/ml by multiplying by 0.142.

In addition, for cohort 2, individuals provided blood for assessment of neutralisation assays. The ability of sera to neutralise SARS-CoV-2 virus was assessed by neutralisation assay on Vero cells. Sera were serially diluted in OptiPRO SFM (Life Technologies) and incubated for 1h at room temperature with 100 TCID50/well of SARS-CoV-2/England/IC19/2020 and transferred to 96-well plates preseeded with Vero-E6 cells. Serum dilutions were performed in duplicate. Plates were incubated at 37°C, 5% CO2 for 42 h before fixing cells in 4% PFA. Cells were treated with methanol 0.6% H2O2 and stained for 1h with a 1:3000 dilution of 40143-R019 rabbit mAb to SARS-CoV-2 nucleocapsid protein (Sino Biological). A 1:3000 dilution of sheep anti-rabbit HRP conjugate (Sigma) was then added for 1 h. TMB substrate (Europa Bioproducts) was added and developed for 20 mins before stopping the reaction with 1M HCl. Plates were read at 450nm and 620nm and the concentration of serum needed to reduce virus signal by 50% was calculated to give NT50 values.

### Performance Analysis

The primary outcome of the study was sensitivity and specificity of each LFIA in detecting SARS-CoV-2 IgG antibodies. Statistical analyses were performed with MedCalc. Graphical analyses was performed using GraphPad Prism 9.1.2 software.

### Ethics and Approvals

Ethics approvals were sought for each cohort prior to commencement of the study. The study was approved by the Health Research Authority, Research Ethics Committee (Reference: 20/WA/0123).

## Results

### Cohort Characteristics

The characteristics of the participants are described in Tables 1 and 2.

**Table 1.** Clinical Characteristics Renal Transplant Cohort.

| Characteristic |  | All patients<br>N=108 (%) | Anti-S<br>Seronegative<br>N= 36 (%) | Anti-S<br>Seropositive<br>N= 72 (%) |
| --- | --- | --- | --- | --- |
| **Gender** | Male | 74 (68.5) | 25 (69.4) | 49 (68.1) |
|  | Female | 34 (31.5) | 11 (30.6) | 23 (31.9) |
| **Age** | Years (Median) | 54 (41-65) | 44 (38-74) | 56 (44-64) |
| **Ethnicity** | White | 52 (48.1) | 17 (47.2) | 35 (48.6) |
|  | Black | 8 (7.4) | 2 (5.6) | 6 (8.3) |
|  | Asian | 34 (31.5) | 11 (30.6) | 23 (31.9) |
|  | Other | 14 (13.0) | 6 (16.7) | 8 (11.1) |
| **Cause of ESKD** | Polycystic kidney disease | 9 (8.3) | 4 (11.1) | 5 (6.9) |
|  | Glomerulonephritis | 41 (38.0) | 12 (33.3) | 29 (40.3) |
|  | Diabetic nephropathy | 18 (16.7) | 7 (19.4) | 11 (15.3) |
|  | Urological | 7 (6.5) | 2 (5.6) | 5 (6.9) |
|  | Unknown | 26 (24.1) | 8 (22.2) | 18 (25.0) |
|  | Other | 7 (6.5) | 3 (8.3) | 4 (5.6) |
| **Vaccinated ≤1 year post transplant** | Yes | 6 (5.6) | 1 (2.8) | 5 (6.9) |
|  | No | 102 (94.4) | 35 (97.2) | 67 (93.1) |
| **Time vaccinated post-transplant** | Years (Median) | 6.3 (2.9-11.9) | 5.7 (2.8-11.7) | 6.5 (2.9-12.0) |
| **Diabetes** | No | 75 (69.4) | 27 (75.0) | 48 (66.7) |
|  | Yes | 33 (30.6) | 9 (25.0) | 24 (33.3) |
| **Vaccine type** | BNT1262b2 | 51 (47.2) | 12 (33.3) | 39 (54.2) |
|  | ChAdOx1 | 57 (52.8) | 24 (66.7) | 33 (45.8) |
| **Time between vaccinations** | Days (median) | 77 (73-80) | 77 (71-80) | 77 (74-80) |
| **Time of serological test post-boost** | Days (median) | 34 (29-38) | 34 (29-36) | 34 (30-38) |
| **Prior COVID exposure** | No | 89 (82.4) | 36 (100.0) | 53 (73.6) |
|  | Yes | 19 (17.6) | - | 19 (26.4) |

**Table 2.** Clinical Characteristics of Healthcare Worker Cohort.

| Characteristic |  | All patients<br>N=40 (%) | Anti S<br>Seronegative |  | Anti S<br>Seropositive |  |
| --- | --- | --- | --- | --- | --- | --- |
|  |  |  | D0<br>N=27<br>(%) | D21<br>N=0<br>(%) | D0<br>N=12<br>(%) | D21<br>N=39<br>(%) |
| **Gender** | Male | 13 (32.5) | 10 (37) | 0 | 3 (25) | 13 (33.3) |
|  | Female | 27 (67.5) | 17 (63) | 0 | 9 (75) | 27 (66.6) |
| **Age** | Years average<br>(range) | 43 (23-71) | 46 | - | 38 | 43 |
| **Vaccine type** | BNT1262b2<br>BioNTech Pfizer | 40 (100) | - | - | - | - |
| **Time between vaccinations** | Days average<br>(range) | 65 (57-92) | - | - | - | - |

### LFIA IgG positivity and antibody titres in serum

The combined results describe both Cohort 1 and Cohort 2 (n=186 samples, Figure 1). The median anti-S titre was 229.5 BAU/ml and mean was 1077 BAU/ml. Of those which scored negative on LFIA (n=68), anti S antibodies were detected in 10 samples, of which 7 had anti-S titre levels <10 BAU/ml (7.82, 8.02, 8.51, 8.78, 9.2, 9.42, 9.72). The other 58 negative LFIA tests had undetectable anti-S levels (<7.10 BAU/ml). Of those samples which scored positive on LFIA (n=118), 4 had undetectable laboratory anti-S levels. The remaining 114 samples had a range of 5670, with a minimum of 9.66 BAU/ml and maximum titre of 5680 BAU/ml. Further investigation of serology is being carried out in some of the patients with detectable antibodies and a negative LFIA result.

**Figure 1.**
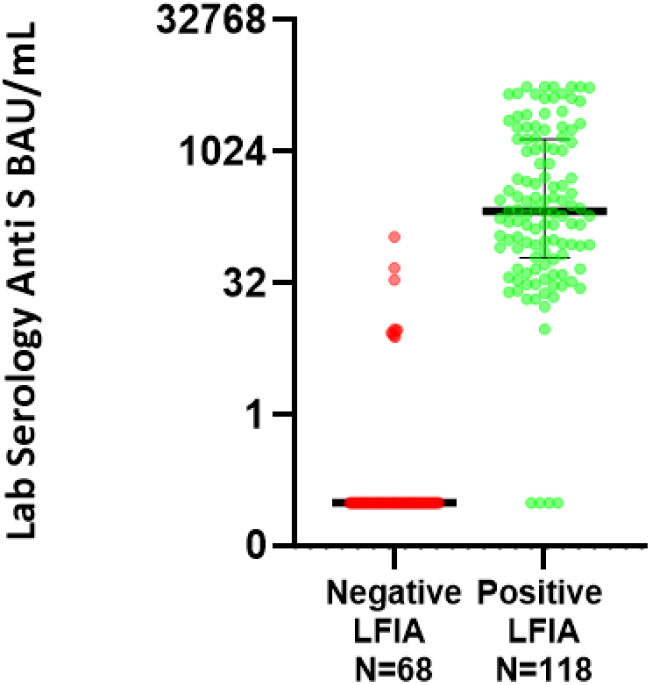
Quantitative anti-S titres (Abbott) between those testing positive and negative on Fortress lateral flow assay for combined cohorts 1 and 2.

### Test sensitivity and specificity

Using the threshold value for positivity on serological testing of ≥7.10 BAU/ml, the overall performance of the test in these combined cohorts (n=186 samples, Figure 1) produce an estimate of sensitivity of 91.94% (95% CI 85.67% to 96.06%) and specificity of 93.55% (95% CI 84.30% to 98.21%) using the Abbott Architect assay as reference standard.

### Live virus neutralisation

Neutralisation titres (NT50) were significantly higher in those with positive LFIA compared to those without (Figure 2A). Only one LFIA-negative sample had detectable neutralisation assay using a threshold for positivity of (NT50 of 15 with an anti-S antibody titre of 7.8 BAU/ml). For individuals with detectable IgG on LFIA only 2/34 did not have significant evidence of viral neutralisation.

**Figure 2.**
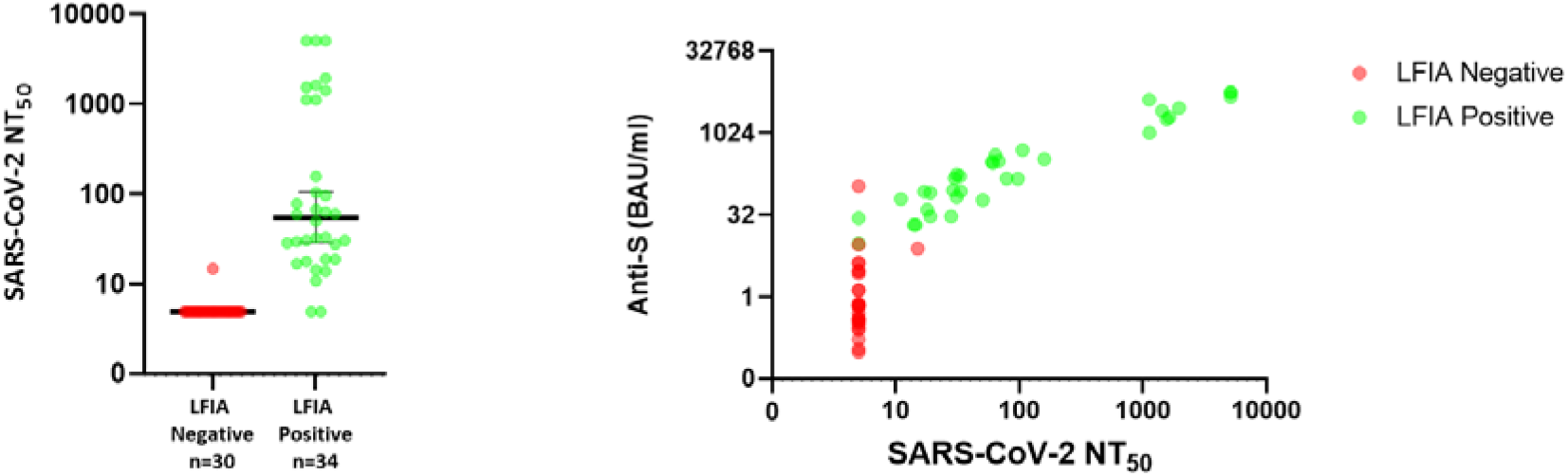
(A) Distribution of NT50 (neutralisation titre) values for individuals within cohort 2 according to whether positive or negative by LFIA (B) Relationship between neutralisation titre and anti-S titre in Abbott assay (green representing those LFIA positive, red LFIA negative).

## Discussion

This study demonstrates that the Fortress LFIA device performs well in detecting IgG antibodies when comparing against a serological assay widely used in NHS services. LFIAs have been a helpful tool the assessment of population antibody prevalence of SARS-CoV-2, and can play a role in informing vaccination strategy going forwards. The Fortress LFIA has been assessed previously for its performance following natural infection (4), though did not meet MHRA criteria for individual use which recommend antibody tests should have a sensitivity of >98% (95% CI 96% to 100%) and specificity of >98% on a minimum of 200 known negative controls (7). The test has undergone evaluation for home self-testing (6) and has since been used widely in community studies of antibody prevalence in England.

The performance of the LFIA in the cohorts of vaccinated individuals here demonstrates slightly higher sensitivity than previously reported for natural infection, though this difference is not significant. This is likely to reflect higher background titres of antibody following vaccination, particularly after second doses, when compared to natural infection in the community at least in the healthcare worker cohort. The LFIA device does not detect very low levels of antibody which may still correlate with protection from severe disease and/ or hospitalisation. However, in the general population, the number of such individuals with low titres following two vaccinations will be low (in contrast to the renal transplant cohort studied here).

A small number of LFIA tests appear to produce false positive results (n=4) with absent antibodies when compared to the Abbott assay. This leads to a slightly lower estimate of specificity, although confidence around this is wide and overlapping with previous estimates. Further work will be undertaken to assess alternative laboratory assays, particularly for comparisons at low levels of antibody.

There is growing evidence that the presence of neutralising antibodies in sera is highly predictive of immune protection from symptomatic Covid-19 disease (8,9). Although it could be argued that the LFIA is a relatively insensitive assay for detecting total Spike specific antibody, the threshold antibody titre required for a positive score is close to the level at which neutralizing antibody can be reliably measured (Figure 2a). This suggests that antibody positivity on the LFIA may give some indication of protection from symptomatic disease and thus could be useful to measure any waning of vaccine induced immunity in different populations that might indicate requirement for administration of booster doses.

In terms of study limitations it is important to note that, although the LFIAs were self-tested in the clinic or vaccination centre, participants had access to support from trained healthcare professionals when required. This does not fully replicate the ‘real-world’ application of LFIAs where users will be following a detailed guide in their own homes. Furthermore, the patient cohort includes healthcare workers and as such may demonstrate a greater affinity for self-testing than members of the general population. However, the primary purpose of this study was to evaluate the diagnostics accuracy of the test.

The performance of the LFIA evaluated is sufficiently good that it can continue to play a helpful role in the assessment of population antibody responses resulting from widespread infection and high levels vaccination coverage, particularly given the correlation of LFIA results with the functional measure of live virus neutralisation. Over time, antibody titres will begin to wane and ongoing population surveillance may have a helpful role in informing decisions on priority groups and timing of booster vaccines.

## Data Availability

Harvard Dataverse Repository: "Replication Data for: SARS-CoV-2 Antibody Lateral Flow Assay for Possible Use in Seroprevalence Surveys: a Diagnostic Accuracy Study", https://doi.org/10.7910/DVN/KCDZIN, This project contains the following underlying data:
- Anti Spike Protein LFIA For HCW Cohort
- Anti Spike Protein LFIA For Renal Cohort
- Neutralisation Titres for HCW Cohort

https://doi.org/10.7910/DVN/KCDZIN

## Data Availability

Harvard Dataverse Repository: “Replication Data for: SARS-CoV-2 Antibody Lateral Flow Assay for Possible Use in Seroprevalence Surveys: a Diagnostic Accuracy Study”, https://doi.org/10.7910/DVN/KCDZIN, This project contains the following underlying data:

- Anti Spike Protein LFIA For HCW Cohort
- Anti Spike Protein LFIA For Renal Cohort
- Neutralisation Titres for HCW Cohort

## Acknowledgements

HW is a National Institute for Health Research (NIHR) Senior Investigator and acknowledges support from NIHR Biomedical Research Centre of Imperial College NHS Trust, NIHR School of Public Health Research, NIHR Applied Research Collaborative North West London, and Wellcome Trust (UNS32973). GC is supported by an NIHR Professorship. AD is an NIHR senior investigator and DA and PE are Emeritus NIHR Senior Investigators. SR acknowledges support from MRC Centre for Global Infectious Disease Analysis, National Institute for Health Research (NIHR) Health Protection Research Unit (HPRU), Wellcome Trust (200861/Z/16/Z, 200187/Z/15/Z), and Centres for Disease Control and Prevention (US, U01CK0005-01-02). PE is Director of the MRC Centre for Environment and Health (MR/L01341X/1, MR/S019669/1). PE acknowledges support from the NIHR Imperial Biomedical Research Centre and the NIHR HPRUs in Chemical and Radiation Threats and Hazards and in Environmental Exposures and Health, the British Heart Foundation Centre for Research Excellence at Imperial College London (RE/18/4/34215), Health Data Research UK (HDR UK) and the UK Dementia Research Institute at Imperial (MC_PC_17114).

## Funding sources

The study was funded by the Department of Health and Social Care in England.

## Conflicts of interest

We declare no competing interests.

## Notes

### Competing Interest Statement

The authors have declared no competing interest.

### Clinical Trial

This study has not been registered on a trial registry.

## References

(1) Maple PAC. Population (Antibody) Testing for COVID-19—Technical Challenges, Application and Relevance, an English Perspective. Vaccines (Basel). 2021; 9 (6): 550. Available from: doi: 10.3390/vaccines9060550 Available from: https://search.proquest.com/docview/2536494082.

(2) Ward H, Atchison C, Whitaker M, Ainslie K, Elliot J, Okell L, et al. Antibody prevalence for SARS-CoV-2 in England following first peak of the pandemic: REACT2 study in 100,000 adults. bioRxiv. 2020. Available from: http://hdl.handle.net/10044/1/81986.

(3) Ward H, Cooke G, Atchison C, Whitaker M, Elliott J, Moshe M, et al. Declining prevalence of antibody positivity to SARS-CoV-2: a community study of 365,000 adults. Cold Spring Harbor Laboratory. 2020. Available from: http://hdl.handle.net/10044/1/83634.

(4) Moshe M, Daunt A, Flower B, Simmons B, Brown JC, Frise R, et al. SARS-CoV-2 lateral flow assays for possible use in national covid-19 seroprevalence surveys (React 2): diagnostic accuracy study. BMJ; 2021.

(5) Flower B, Brown JC, Simmons B, Moshe M, Frise R, Penn R, et al. Clinical and laboratory evaluation of SARS-CoV-2 lateral flow assays for use in a national COVID-19 seroprevalence survey. BMJ; 2020.

(6) Atchison C, Pristerà P, Cooper E, Papageorgiou V, Redd R, Piggin M, et al. Usability and acceptability of home-based self-testing for SARS-CoV-2 antibodies for population surveillance.

(7) MHRA. Target product profile antibody tests to help determine if people have immunity to SARS-CoV-2. Available from: https://assets.publishing.service.gov.uk/ government/uploads/system/uploads/attachment_data/file/883897/Target_Product_ Profile_antibody_tests_to_help_determine_if_people_have_immunity_to_SARSCoV-2_Version_2.pdf [Accessed 21.6.21].

(8) Earle KA, Ambrosino DM, Fiore-Gartland A, Goldblatt D, Gilbert PB, Siber GR, et al. Evidence for antibody as a protective correlate for COVID-19 vaccines. Cold Spring Harbor Laboratory; 2021.

(9) Khoury DS, Cromer D, Reynaldi A, Schlub TE, Wheatley AK, Juno JA, et al. Neutralizing antibody levels are highly predictive of immune protection from symptomatic SARS-CoV-2 infection. Springer Science and Business Media LLC; 2021.

