## Supplemental for "SARS-CoV-2 Antibody Lateral Flow Assay for antibody prevalence studies following vaccine roll out: a Diagnostic Accuracy Study"

**Renal Transplant Cohort Test Performance**

Of those samples which scored positive on LFIA (n=69), 3 had undetectable laboratory anti-S levels. The remaining 66 samples had a range of 5563, with maximum titre level of 5680 BAU/ml and minimum of 17.46 BAU/ml. The median anti-S titre of the true positive LFIAs was 281.6 BAU/ml and mean was 1177 BAU/ml. Of those which scored negative on LFIA (n=39), anti S antibodies were detected in six samples of which 4 had anti-S titre levels <10 BAU/ml (8.02, 8.51, 9.42 and 9.72). The other 33 negative LFIA samples had undetectable anti-S levels (<7.10 BAU/ml). 3 individuals with negative laboratory serology despite positive LFIAs are being further investigated.

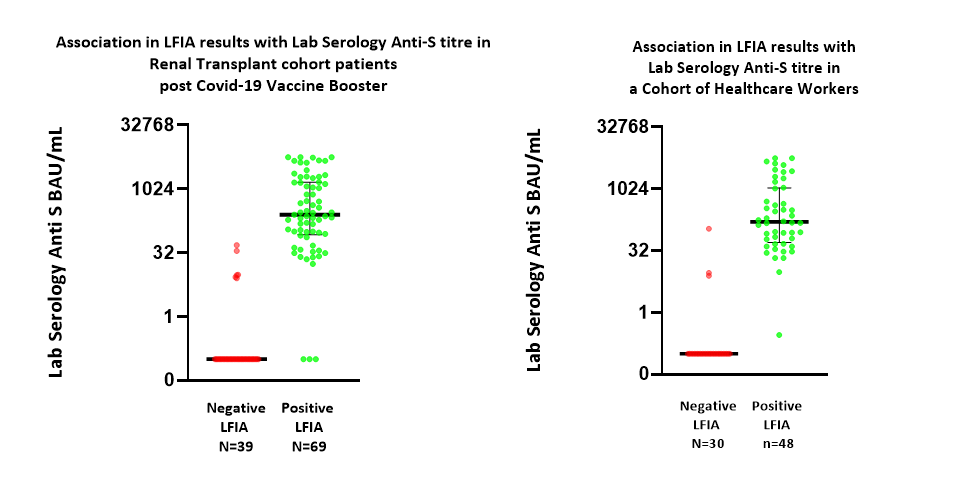

**Figure 2.** Quantitative anti-S titres (Abbott) between those testing positive and negative on Fortress lateral flow assay for renal transplant cohort.

**Healthcare Worker Cohort Test Performance**

Of those samples which scored positive on LFIA (n=49), 1 had undetectable anti-S levels. The remaining 48 samples had a range of 5670, with maximum titre level of 5680 BAU/ml and minimum of 9.66 BAU/ml. The median anti-S titre of true positives was 166.9 BAU/ml and mean was 957.4 BAU/ml. Of those which scored negative on LFIA (n=29), anti-S antibodies were detected in four samples, three of which were <10 BAU/ml (7.82, 8.78 and 9.2). The other 25 samples had anti-S levels <7.10ml.

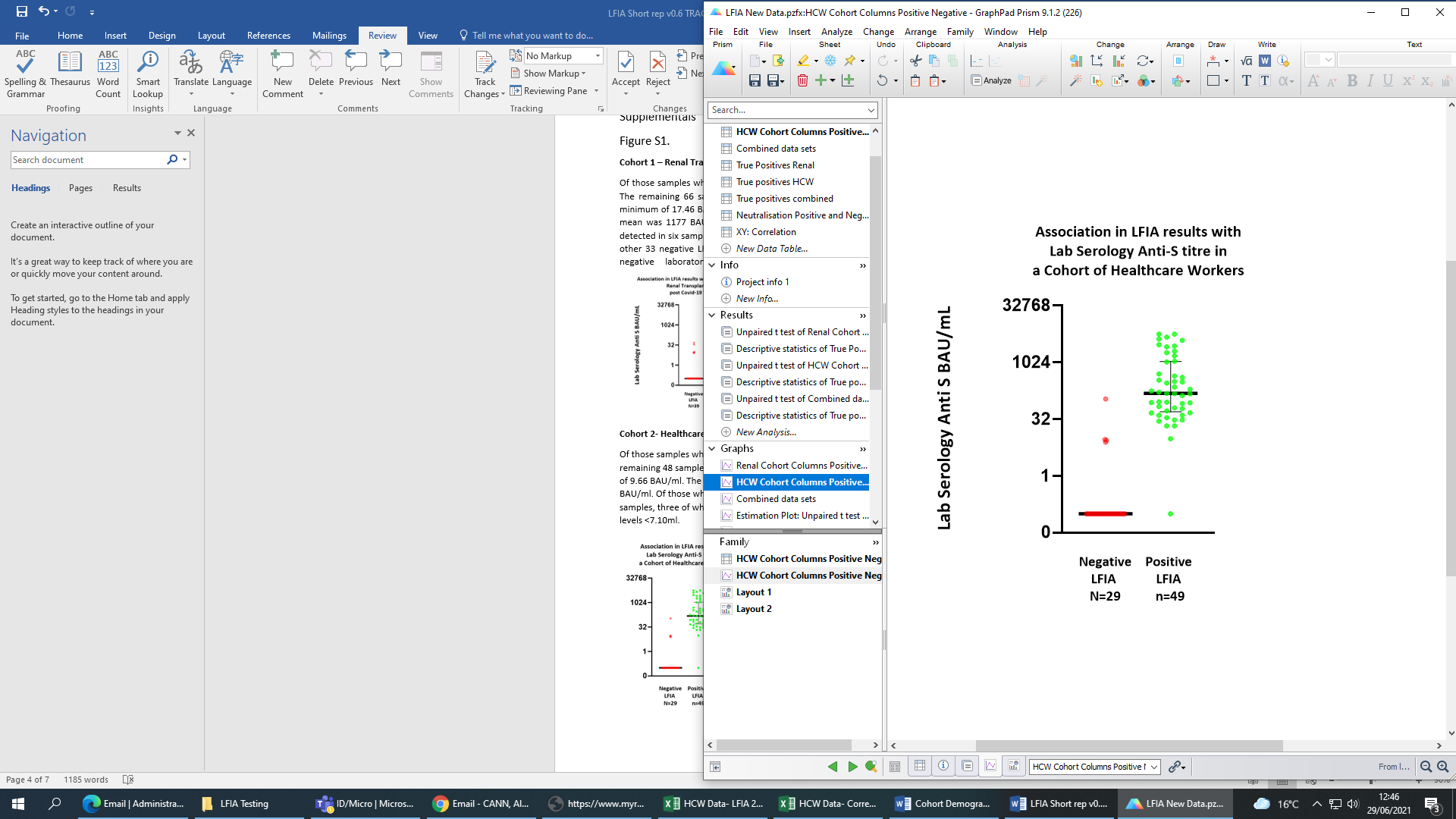

**Figure 3.** Quantitative anti-S titres (Abbott) between those testing positive and negative on Fortress lateral flow assay for healthcare worker cohort.
